# Synaptic loss in behavioural variant frontotemporal dementia revealed by [^11^C]UCB-J PET

**DOI:** 10.1101/2022.01.30.22270123

**Authors:** Maura Malpetti, P. Simon Jones, Thomas E. Cope, Negin Holland, Michelle Naessens, Matthew A. Rouse, George Savulich, Tim D. Fryer, Young T. Hong, Selena Milicevic Sephton, Franklin I. Aigbirhio, John T. O’Brien, James B. Rowe

## Abstract

Synaptic loss is an early feature of neurodegenerative disease models, and is often severe in *post mortem* clinical studies, including frontotemporal dementia. Positron emission tomography (PET) imaging with radiotracers that bind to synaptic vesicle glycoprotein 2A enables quantification of synapses *in vivo*. This study used [^11^C]UCB-J PET in people with behavioural variant frontotemporal dementia (bvFTD), testing the hypothesis that synaptic loss is severe and related to clinical severity. We performed a cross-sectional observational study of bvFTD, *versus* healthy controls, in which participants underwent neurological examination, neuropsychological assessment, magnetic resonance imaging (MRI) and [^11^C]UCB-J PET. Patients were recruited from the Cambridge Centre for Frontotemporal Dementia at the University of Cambridge, and healthy volunteers from the UK National Institute for Health Research Join Dementia Research register. Eleven people with a clinical diagnosis of probable bvFTD and 25 age- and sex-matched healthy controls were included. All participants underwent dynamic [^11^C]UCB-J PET imaging, structural MRI and a neuropsychological battery, including the Addenbrooke’s cognitive examination (ACE-R), and INECO frontal screening (IFS). General linear models were used to compare [^11^C]UCB-J binding potential maps between groups, and correlate synaptic density with cognitive performance and clinical features in patients. Group-comparison and correlation analyses were also performed using partial-volume corrected [^11^C]UCB-J binding potential from regions of interest (ROIs). Patients with bvFTD showed severe synaptic loss compared to controls. In particular, [^11^C]UCB-J binding was significantly reduced bilaterally in medial and dorsolateral frontal regions, inferior frontal gyri, anterior and posterior cingulate gyrus, insular cortex and medial temporal lobe. Synaptic loss in the left frontal and cingulate regions correlated significantly with cognitive impairments as assessed with ACE-R and IFS. Results from ROI-based analyses mirrored the voxel-wise results. In keeping with preclinical models, and human post mortem data, there is widespread frontotemporal loss of synapses in symptomatic bvFTD, in proportion to disease severity. [^11^C]UCB-J PET could support translational studies and experimental medicines strategies for new disease-modifying treatments for neurodegeneration.

## Introduction

Frontotemporal dementia is a clinically and pathologically heterogeneous group of neurodegenerative conditions. Similar clinical phenotypes arise from different pathologies (Bang *et al*., 2015; Olney *et al*., 2017), including 3R or 4R tauopathy, and TDP-43 pathology. Despite this molecular heterogeneity, *post mortem* human studies (Lipton *et al*., 2001; Clare *et al*., 2010) and animal models (Yoshiyama *et al*., 2007) suggest that the spectrum of frontotemporal dementia is characterised by early and severe synaptic loss, even preceding neuronal death and atrophy. Our hypothesis was that in people with frontotemporal dementia, synaptic loss is severe and proportionate to clinical severity. Regionally specific and clinically relevant loss of synapses has been shown in the tauopathies of Alzheimer’s disease (Chen *et al*., 2018; Mecca *et al*., 2020; Vanhaute *et al*., 2020; Coomans *et al*., 2021; O’Dell *et al*., 2021), progressive supranuclear palsy and corticobasal degeneration (Holland *et al*., 2020, 2021). TDP-43 related frontotemporal lobar degeneration is also associated with synaptic loss (Ni *et al*., 2021; Riku *et al*., 2021). For the behavioural variant of frontotemporal dementia (bvFTD), characterised by progressive personality and behaviour changes, and cognitive decline, including executive impairments (Piguet *et al*., 2011; Rascovsky *et al*., 2011), we propose that synaptic loss would be most prominent in frontotemporal regions.

Previous evidence for synaptic loss in frontotemporal dementia has been indirect, including reduced synaptic density in carriers of mutations associated with frontotemporal dementia (Malpetti *et al*., 2021), abnormal synaptic markers in cerebrospinal fluid (Goetzl *et al*., 2016; Van Der Ende *et al*., 2020; Bergström *et al*., 2021), and progressive frontotemporal hypometabolism that is disproportionate to atrophy, indicated by [^18^F]FDG PET (Diehl-Schmid *et al*., 2007; Malpetti *et al*., 2019; Bejanin *et al*., 2020; see Chételat *et al*., 2020 for review). Recently, new tools to quantify synaptic density *in vivo* have been developed, including radioligands that bind selectively to synaptic vesicle protein 2A (SV2A) as an assay of synaptic density (Finnema *et al*., 2016; Heurling *et al*., 2019). Synaptic density changes only slightly in mid- and later-life (Finnema *et al*., 2016; Michiels *et al*., 2021), providing a stable background against which to assess the effect of disease and disease-severity.

Here, we used PET with the SV2A radioligand [^11^C]UCB-J to assess the regional distribution of synaptic loss in patients with a clinical diagnosis of probable bvFTD, compared to age- and sex-matched controls. Synaptic density was estimated using non-displaceable binding potential (BP_ND_), a metric proportional to binding site density. In this context, if proven useful, [11C]UCB-J PET could become an important tool for patient stratification and a surrogate endpoint in clinical trials and early interventions. PET measures for synaptic density would give additive and complementary information to fluid marker evidence, which do not elucidate *in vivo* spatial distributions of brain changes and may be less sensitive in prodromal stages. This allows to test the utility of [^11^C]UCB-J as an *in vivo* marker for synaptic loss in frontotemporal dementia, irrespective of whether the underlying pathology is Tau or TDP-43. We predicted substantial synaptic loss in frontotemporal regions across all patients, and a correlation with clinical severity.

## Materials and Methods

### Participants

Eleven patients with a clinical diagnosis of probable bvFTD (Rascovsky *et al*., 2011) were recruited from the Cambridge Centre for Frontotemporal Dementia at Cambridge University Hospitals NHS Foundation Trust and the University of Cambridge. Twenty-five healthy volunteers were recruited from the UK National Institute for Health Research Join Dementia Research (JDR) register.

Participants were screened for exclusion criteria: current or recent history (within the last 5 years) of cancer, current use of the anti-convulsant medication levetiracetam (that binds to SV2A, the target of [^11^C]UCB-J), history or MRI evidence of ischaemic or haemorrhagic stroke, any severe physical illness or co-morbidity that would limit the ability to fully and safely participate in the study, and any contraindications to MRI. Eligible volunteers underwent a clinical and neuropsychological assessment including the Mini-mental State Exam (MMSE), revised Addenbrooke’s Cognitive Examination (ACE-R), INECO frontal screening (IFS), the Frontotemporal Dementia Rating Scale (FTD-RS), and the Clinical Dementia Rating Scale (CDR). Within 3 months, all participants underwent brain imaging with 3T MRI and PET scanning with [^11^C]UCB-J ((*R*)-1-((3-(methyl-^11^C)pyridin-4-yl)methyl)-4-(3,4,5-trifluorophenyl)pyr-rolidin-2-one).

The research protocol was approved by the Cambridge Research Ethics Committee (18/EE/0059) and the Administration of Radioactive Substances Advisory Committee. All participants provided written informed consent in accordance with the Declaration of Helsinki.

### Imaging acquisition and processing

Full details of the protocol for [^11^C]UCB-J synthesis, data acquisition, image reconstruction and kinetic analysis have been published elsewhere (Holland *et al*., 2020; Milicevic Sephton *et al*., 2020; Malpetti *et al*., 2021). In brief, dynamic PET data acquisition was performed on a GE SIGNA PET/MR (GE Healthcare, Waukesha, USA) for 90 minutes following [^11^C]UCB-J injection, with attenuation correction including the use of a multi-subject atlas method (Burgos *et al*., 2014) and improvements to the brain MRI coil component (Manavaki *et al*., 2019). Each emission image series was aligned and rigidly registered to T1-weighted MRI acquired in the same session (TR=3.6 msec, TE=9.2 msec, 192 sagittal slices, in-plane voxel dimensions 0.55×0.55 mm (subsequently interpolated to 1.0×1.0 mm); slice thickness 1.0 mm).

For each subject a [^11^C]UCB-J BP_ND_ map was determined from dynamic images corrected for partial volume effects at the voxel level using the iterative Yang method (Erlandsson *et al*., 2012). BP_ND_ was calculated using a basis function implementation of the simplified reference tissue model, with centrum semiovale as the reference tissue (Rossano *et al*., 2019). Each BP_ND_ map was warped to the ICBM 152 2009a asymmetric MR template using parameters from the spatial normalisation of the co-registered T1 MR image with Advanced Normalization Tools (ANTs; http://www.picsl.upenn.edu/ANTS/). For voxel-wise analyses, the spatially normalised [^11^C]UCB-J BP_ND_ maps were spatially smoothed with a 10 mm full width at half maximum (FWHM) gaussian kernel prior to statistical analysis.

For regional analysis, we used the n30r83 Hammers atlas (http://brain-development.org) modified to include segmentation of brainstem and cerebellum, and non-rigidly registered to the T1-weighted MRI of each participant. Regions were multiplied by a binary grey matter mask constructed in Statistical Parametric Mapping (SPM12 v7771, Institute of Neurology, London, UK) based on a >50% probability map smoothed to PET spatial resolution, and geometric transfer matrix partial volume correction was applied to each image of the dynamic series (Rousset *et al*., 1998). Regional BP_ND_ was determined with the same kinetic modeling approach and reference tissue as for the BP_ND_ maps.

### Statistical analyses

Voxel-wise analyses were performed using SPM12. First, a two-sample t-test was performed to compare the bvFTD group to controls across the whole brain. Second, to test the association between synaptic density and clinical severity in people with bvFTD, voxel-wise general linear models were applied across all patients, including ACE-R and IFS scores as dependent variables in separate models (for IFS, the analyses used 10 patients due to missing data). Similar models were applied to test for associations between [^11^C]UCB-J BP_ND_, age and symptom duration. All results were tested at an uncorrected voxel height threshold of p < 0.001 combined with a familywise error (FWE) corrected cluster threshold of p < 0.05; peak voxels in clusters that reached the conservative voxel-level familywise error threshold (FWE p <0.05) are also indicated.

We performed complementary analyses on regions of interest, using R version 4.0.0 (R Core team 2020, https://www.r-project.org/) and JASP (JASP team: https://jasp-stats.org/). [^11^C]UCB-J BP_ND_ values from regions of interest were aggregated into left and right frontal lobe, temporal lobe, parietal lobe, occipital lobe, cingulate cortex, and included in the analyses alongside insula cortex, hippocampus, amygdala, and thalamus. First, two-sample t-tests were performed in each region to compare the bvFTD group to controls. Across all brain regions, p-values from t-tests were corrected with false-discovery rate (FDR) correction, and Cohen’s d effect sizes were calculated. As explorative analyses, we also performed two-sample t-tests on [^11^C]UCB-J BP_ND_ values of bvFTD group *vs* controls considering smaller regions, obtained from the Hammers atlas with and without partial-volume correction. Second, Spearman’s correlation analyses were performed to test the association between cognitive performance and [^11^C]UCB-J BP_ND_ in cortical regions. In addition to the above frequentist statistics, we also present Bayesian analogous tests for regional effects (https://osf.io/gny35/), furnishing the strength of the evidence for the null *vs* alternate hypotheses.

## Data Availability Statement

Anonymized data may be shared upon request to the corresponding or senior author from a qualified investigator for non-commercial use, subject to restrictions according to participant consent and data protection legislation.

## Results

### Descriptive statistics

Demographic and clinical features for the two groups are given in Table 1. The groups were similar in age and sex, with cognitive deficits among patients typical for bvFTD.

**Table 1.** Demographic and clinical characteristics for control and bvFTD groups. Mean and standard deviation are reported for each continuous variable. Group comparisons were performed with two-sample t-tests for continuous variables, and chi-square test for sex.

|  | **Controls**<br>**N=25** | **bvFTD**<br>**N=11** | *Difference* |
| --- | --- | --- | --- |
| Age | 70.2 (7.0) | 65.7 (9.3) | n.s. |
| Sex (F:M) | 9:16 | 2:9 | n.s. |
| MMSE [/30] | 29.5 (1.1) | 22.3 (8.6) | T(34)=2.8, p=0.019, d=1.2 |
| ACE-R [/100] | 96.2 (2.5) | 63.0 (26.9) | T(34)=4.1, p=0.002, d=1.7 |
| Att/Orient [/18] | 17.9 (0.3) | 13.0 (5.6) | T(34)=2.9, p=0.016, d=1.2 |
| Memory [/26] | 24.7 (1.7) | 14.5 (8.4) | T(34)=4.0, p=0.002, d=1.7 |
| Fluency [/14] | 12.4 (1.5) | 4.4 (4.5) | T(34)=5.8, p<0.001, d=2.4 |
| Language [/26] | 25.6 (0.8) | 19.1 (7.6) | T(34)=2.8, p=0.018, d=1.2 |
| Visuospatial [/16] | 15.6 (0.6) | 12.1 (4.0) | T(34)=2.9, p=0.015, d=1.2 |
| IFS | 25.7 (2.4) | 9.4 (7.1) | T(32)=7.0, p<0.001, d=3.1 |
| IFS WM | 7.6 (1.3) | 3.7 (2.4) | T(32)=5.0, p<0.001, d=2.1 |
| Symptom duration (months) | - | 79.9 (40.6) | - |
| FTD-RS % | - | 18.9 (18.9) | - |
| FTD-RS Logit | - | -2.6 (1.7) | - |

Abbreviations: bvFTD = behavioural variant frontotemporal dementia; MMSE = mini-mental state examination; ACE-R = Addenbrooke's cognitive examination revised; Att/orient = attention/orientation score; IFS = INECO frontal screening; WM = working memory; FTD-RS = frontotemporal dementia rating scale; ns = not significant; d = Cohen's d.

### Voxel-wise group comparisons and correlations with cognitive scores

Single-subject [^11^C]UCB-J BP_ND_ maps for individual patients and the average BP_ND_ map across all controls are reported in Figure 1. People with bvFTD showed mild (patient 1) to severe (patient 11) regional synaptic loss compared to controls.

**Figure 1.**
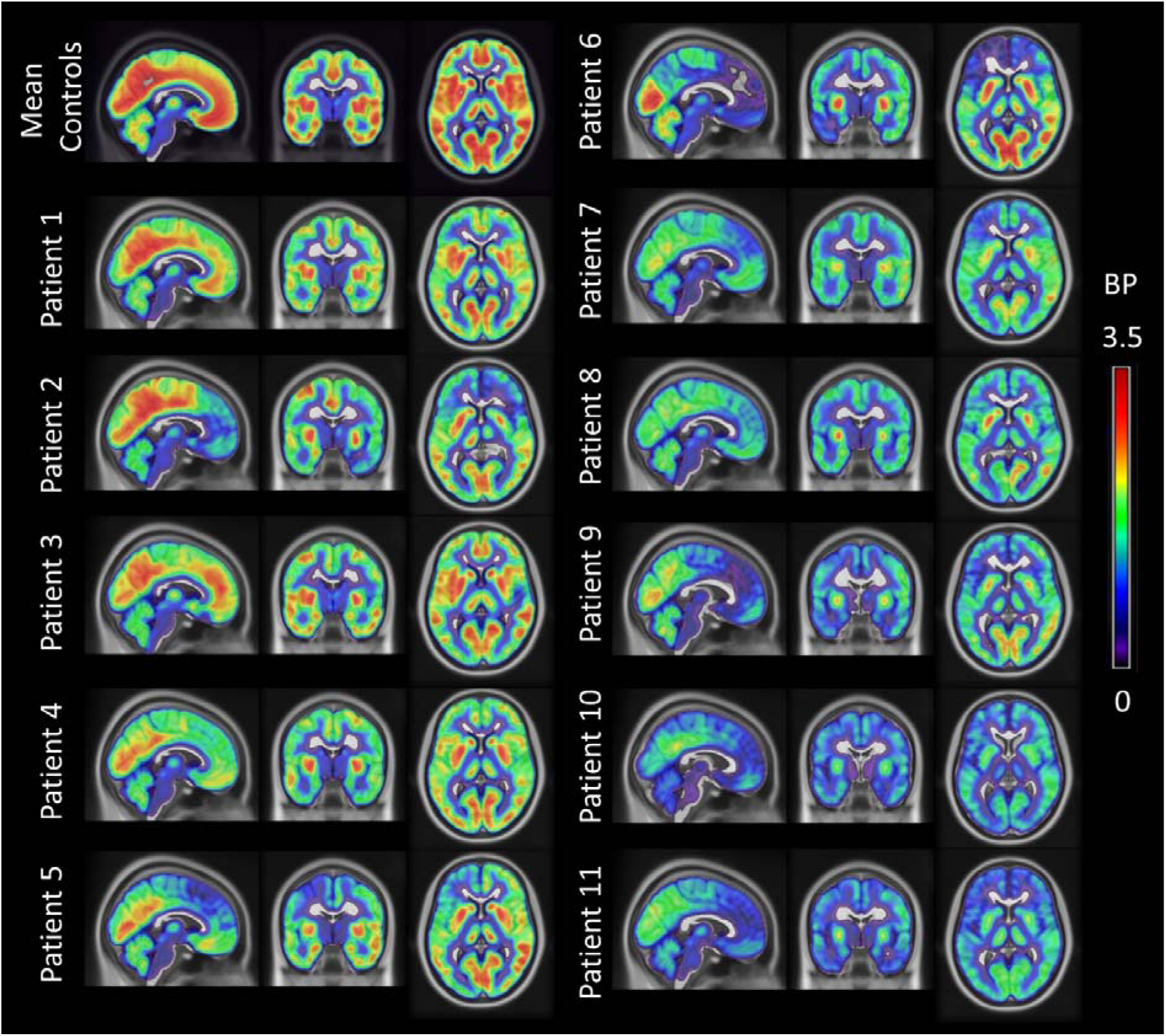
[^11^C]UCB-J binding potential (BP_ND_) maps for each bvFTD patient. The BP_ND_ maps were spatially normalised to ICBM 152 2009a space, masked and smoothed (isotropic 6mm full width at half maximum Gaussian for visualisation). The BP_ND_ maps are overlaid on the ICBM 152 2009a T1 MR template and the slices are reported in the neurological display convention (left on the left). For comparison, the first row on the left shows the corresponding average BP_ND_ map across 25 controls.

At the FWE-corrected voxel-level threshold (p<0.05), the two-sample t-test on [^11^C]UCB-J BP_ND_ maps revealed synaptic loss in the bvFTD group compared to controls (5.1 ≤ t ≤ 9.3) in frontal, temporal, insula and cingulate cortex, bilaterally (Figure 2, panel A). Group differences were most pronounced in medial frontal and orbitofrontal cortex, superior frontal regions, inferior frontal gyrus, insula cortex, anterior and superior temporal lobe, and anterior cingulate cortex (t ≥ 6.4).

**Figure 2.**
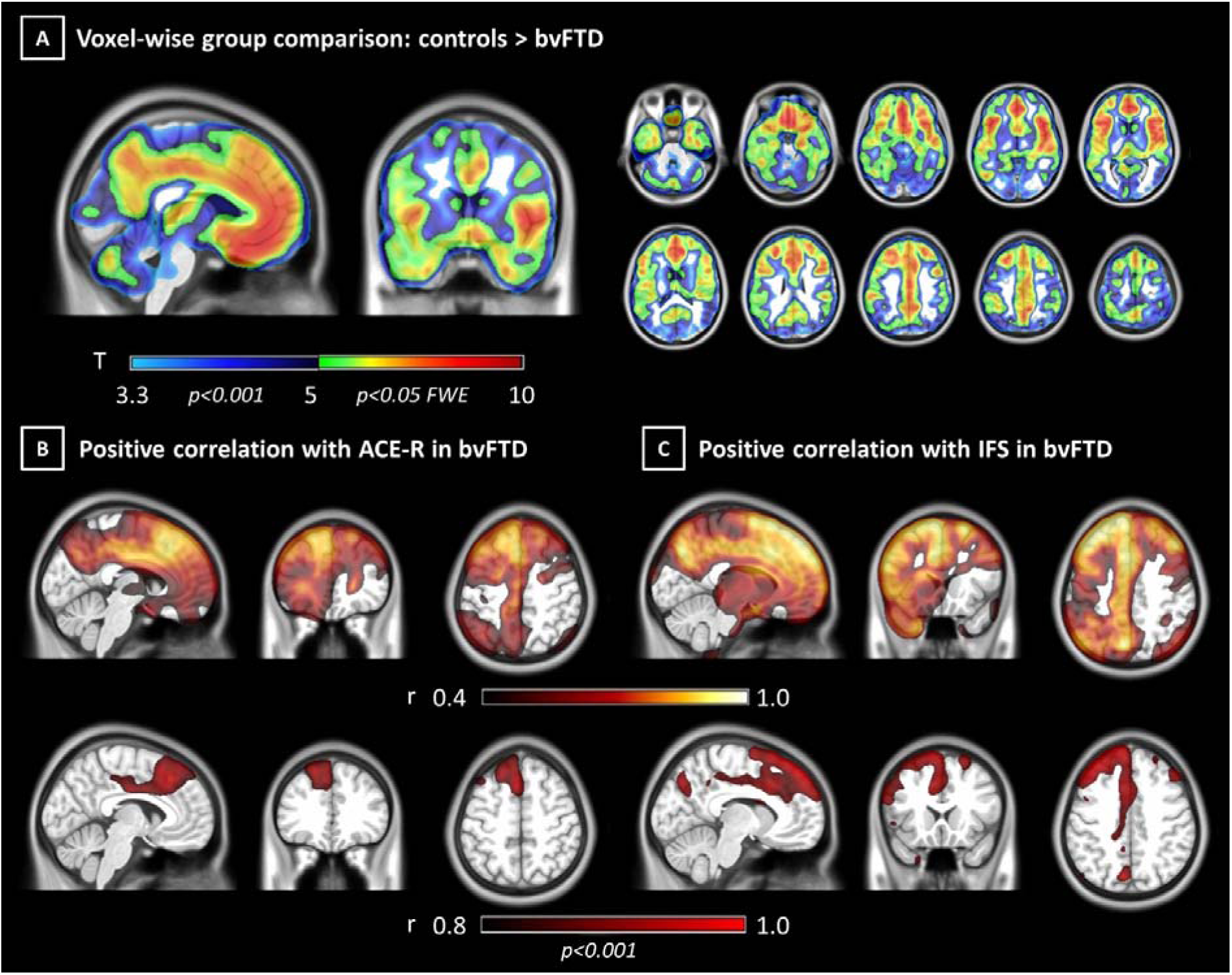
Voxel-wise synaptic loss and association with cognitive impairments in bvFTD. Panel A: t maps from voxel-wise analysis showing higher [^11^C]UCB-J binding potential in controls compared to bvFTD (p < 0.05 FWE-corrected at voxel level). Panels B and C: Coefficient of correlation maps for the bvFTD group between voxel-wise [^11^C]UCB-J binding potential and cognitive performance at ACE-R and IFS (p < 0.001 uncorrected at voxel level, p < 0.05 FWE-corrected at cluster level).

For voxel-wise correlation analyses between [^11^C]UCB-J BP_ND_ and cognition, ACE-R showed a strong positive association (r ≥ 0.8; 6.3 ≤ t ≤ 6.8, p < 0.001 uncorrected at the voxel level and p < 0.05 FWE-corrected at the cluster level) between cognition and [^11^C]UCB-J BP_ND_ in the anterior cingulate gyrus bilaterally, in the left superior and middle frontal gyri, and the medial anterior temporal lobe (Figure 2, panel B). IFS scores correlated positively (r ≥ 0.8; 7.9 ≤ t ≤ 9.9, p < 0.001 uncorrected at voxel level and p < 0.05 FWE-corrected at cluster level) with [^11^C]UCB-J BP_ND_ in the superior and middle frontal gyri bilaterally, left cingulate cortex and inferior frontal gyrus, and left superior parietal gyrus and anterior medial temporal lobe (Figure 2, panel C). Voxel-wise correlation analyses did not reveal significant associations between [^11^C]UCB-J BP_ND_, age and symptom duration (p<0.001 uncorrected).

### ROI-based group comparisons and correlations with cognitive scores

Group comparison results by two-sample t-tests in each region of interest are shown in Figure 3 and in more detail in Supplementary Table 1. bvFTD caused severe synaptic loss in frontal and temporal lobes bilaterally, with the right cingulate and insula cortex showing the most severe loss (−6.9 ≤ t ≤ - 5.6; -2.7 ≤ Cohen’s d ≤ -2.2). Results from explorative two-sample t-tests on sub-regions are reported in Supplemental Material, including partial-volume corrected (Supplementary Table 2) and uncorrected (Supplementary Table 3) regional BP_ND_.

**Figure 3.**
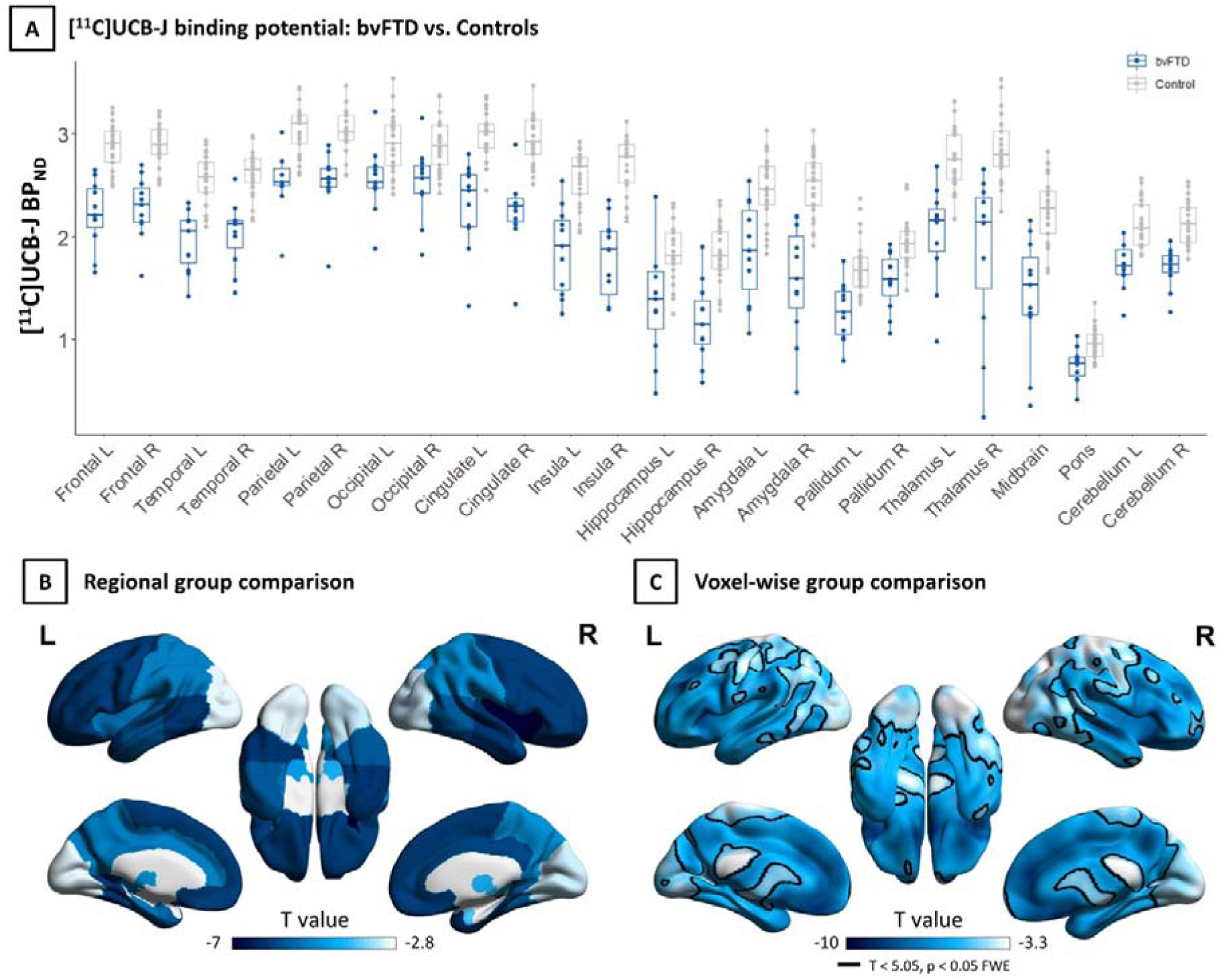
Regional partial-volume corrected binding potential (BP_ND_) values and group comparisons. Panel A: Regional [^11^C]UCB-J BP_ND_ values for bvFTD patients (blue) and controls (grey). Panel B: Regional t values for comparison of [^11^C]UCB-J BP_ND_ between bvFTD patients and controls (p < 0.05 FDR-corrected) surface rendered on. Panel C: For visual comparison with panel B, corresponding t-values obtained from voxel-wise group comparison.

Considering cortical regions of interest, Spearman’s correlation between partial-volume corrected [^11^C]UCB-J BP_ND_ and ACE-R scores revealed strong positive associations in the left frontal lobe (r=0.791, p = 0.002, Figure 4A), cingulate cortex (r=0.700, p = 0.008, Figure 4C) and parietal lobe (r=0.591, p = 0.028, Figure 4E). Spearman correlation analyses with IFS scores showed positive associations with [^11^C]UCB-J BP_ND_ in the same regions: the left frontal lobe (r=0.754, p = 0.006, Figure 4B), cingulate cortex (r=0.620, p = 0.028, Figure 4D) and parietal lobe (r=0.632, p = 0.025, Figure 4F). Similar results were obtained with regional BP_ND_ values without partial volume correction.

**Figure 4.**
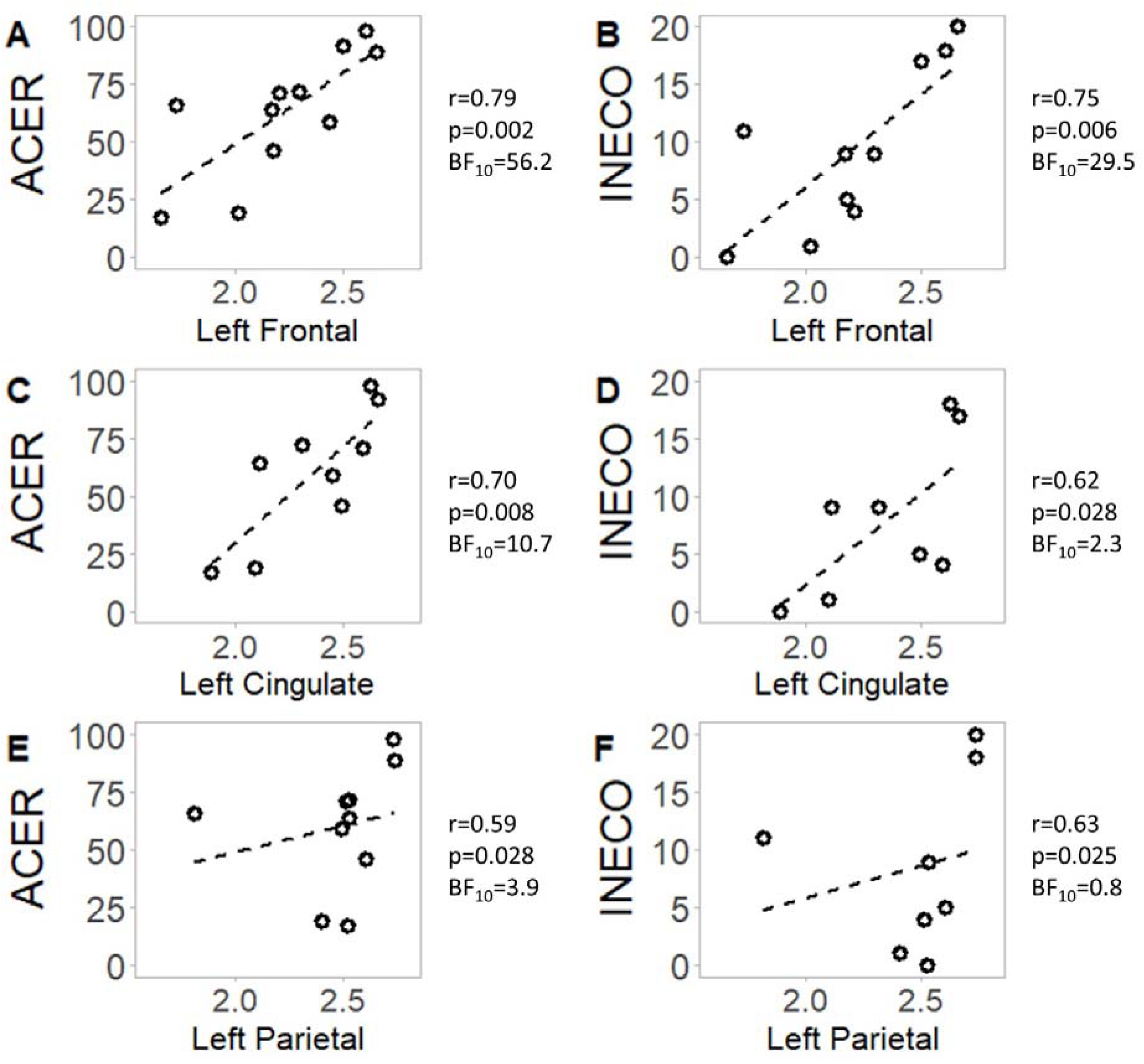
Correlations between regional synaptic density and cognitive performance. [^11^C]UCB-J binding potential in a cortical region is reported on the x axis, while the y axis represents a cognitive test score. Abbreviations: ACER = Addenbrooke’s cognitive examination revised; BF = Bayes factor; IFS = INECO frontal screening; r = Spearman rho.

Bayesian correlation analyses indicated strong evidence for the associations between [^11^C]UCB-J BP_ND_ and ACE-R scores in the left frontal lobe (Bayes Factor (BF) = 56.2), cingulate cortex (BF=10.7), with positive evidence in the parietal lobe (BF=3.9). Bayesian correlation analyses of IFS indicated strong evidence for positive correlation in the left frontal lobe (BF=23.4) but a lack of positive evidence for or against a correlation in the left cingulate cortex (BF=2.3) and parietal lobe (BF=0.8). In Supplementary Table 4, explorative frequentist and Bayesian correlation analyses of regional [^11^C]UCB-J BP_ND_ with MMSE, ACE-R and IFS sub-scores, and FTD-RS are reported.

## Discussion

This study confirms the hypothesis that people with a clinical diagnosis of bvFTD have severe and widespread synaptic loss over frontotemporal cortex, including cingulum and insula. The synaptic density in frontal and cingulate regions as quantified with [^11^C]UCB-J BP_ND_ correlates with patients’ cognitive performance. [^11^C]UCB-J BP_ND_ was not related to the estimated symptom duration or age, in line with previous evidence (Michiels *et al*., 2021). These principal results were observed with and without partial volume correction, and using both voxel-wise and regional analysis. The loss of synapses detected by [^11^C]UCB-J PET raises three points of particular importance.

First, the decline of synaptic health – including synaptic density and synaptic plasticity – will directly affect brain connectivity and learning. Synaptic mediation of neurophysiological connectivity underpins cognitive function, while synaptic plasticity is one of the major determinants of learning and memory in multiple cognitive systems. In Alzheimer’s disease, synaptic loss is more closely related to cognitive impairment than tau burden, beta-amyloid burden or neuronal loss (Spires-Jones and Hyman, 2014). Such a preeminent relationship may also hold in frontotemporal dementia. The distinctive distribution of synaptic loss in bvFTD across frontotemporal, insula and cingulate cortex is consistent with the typical distribution of its molecular pathologies, and its neurocognitive and behavioural profile. In this study, all of our cases were symptomatic (CDR ≥ 1), although we note that behavioural and cognitive changes can emerge in the presymptomatic and prodromal stages of bvFTD, many years before dementia and diagnosis (Rohrer *et al*., 2015; Malpetti *et al*., 2020; Staffaroni *et al*., 2020); and synaptic loss has been identified in the presymptomatic stage of those with highly penetrant mutations such as C9orf72 expansions (Malpetti *et al*., 2021). Early synaptic dysfunction in these regions may explain the subtle pre-symptomatic behavioural change and executive dysfunction. In addition, synaptic loss may determine the neurophysiological signatures of bvFTD as assayed by magnetoencephalography (Hughes *et al*., 2018; Sami *et al*., 2018; Adams *et al*., 2021; Shaw *et al*., 2021; Copet *et al*., 2022) as in Alzheimer’s disease (Coomans *et al*., 2021).

Second, quantifying the degree and distribution of synaptic loss can enrich models of frontotemporal dementia pathogenesis, in preclinical and clinical settings. For example, in preclinical tauopathy models, connectivity more than proximity has been shown to influence the spread of diverse toxic oligomeric tau species (Clavaguera *et al*., 2009, 2013; de Calignon *et al*., 2012; Iba *et al*., 2013; Ahmed *et al*., 2014; Mudher *et al*., 2017). Several mechanisms have been demonstrated at the synapse by which such toxic oligomeric Tau can be transferred between neurons. The oligomeric tau is in turn synaptotoxic (Lasagna-Reeves *et al*., 2011; Sydow *et al*., 2011), and affects synaptic health even in the absence of cell death. In Alzheimer’s disease, *in vivo* human PET and CSF studies recently demonstrated a strong association between tau pathology and reduced synaptic integrity (Vanhaute *et al*., 2020; Casaletto *et al*., 2021; Coomans *et al*., 2021). In other clinical disorders caused by tau-mediated frontotemporal lobar degeneration, the tauopathy seems to be synaptotoxic (Holland *et al*., 2021), whilst synaptic density and connectivity seem to confer vulnerability to molecular pathology (Cope *et al*., 2018; Holland *et al*., 2021). In preclinical models, [^11^C]UCB-J may now be used in autoradiography or *in vivo* PET (Finnema *et al*., 2016; Nabulsi *et al*., 2016; Bertoglio *et al*., 2020; Varnäs *et al*., 2020; Thomsen *et al*., 2021; Xiong *et al*., 2021) to quantify synapse density, where it is highly correlated with better established markers such as synaptophysin (Finnema *et al*., 2016). [^11^C]UCB-J therefore offers a bridge between preclinical and clinical models, where quantitative whole-brain models of pathogenesis in dementia, including frontotemporal dementia, can be improved by the inclusion of synaptic density estimates in addition to structural connectivity (Raj *et al*., 2012) and post-synaptic structural integrity (Mak *et al*., 2021). In addition, it is possible that tau and TDP-43 mediated bvFTD have distinct spatiotemporal profiles of synaptic loss. This could be tested with studies using [^11^C]UCB-J PET in preclinical models and people with genetically determined familial frontotemporal dementia (Van Der Ende *et al*., 2020; Bergström *et al*., 2021), or in due course by *post mortem* examination of those who have undergone PET. However, we suggest that synaptic loss is a mechanistic convergence point, present across diverse molecular pathologies.

Third, with a critical role for synaptic health in mediating between the molecular pathology and cognitive-physiological impairment of frontotemporal dementia, the ability to quantify the degree and distribution of synaptic loss can enhance novel experimental medicine studies (Cope *et al*., 2021). The degree and distribution of loss may be used either for stratification in inclusion or analytical stages of an early-phase clinical trial. It may also be considered as an intermediate marker of efficacy, particularly where the mechanisms of action of a drug is upstream of synaptic loss or is directly related to synaptic resilience and preservation.

[^11^C]UCB-J is not the only potential marker of synaptic health in frontotemporal dementia, and related disorders. A previous study using [^18^F]UCB-H PET to assess synaptic loss did not identify a deficit in frontotemporal dementia *vs* controls, nor was there a difference between frontotemporal dementia and Alzheimer’s disease (Salmon *et al*., 2021). The lower specific binding of [^18^F]UCB-H compared to [^11^C]UCB-J may have contributed to the null result. Results with the alternative ligand [^18^F]-SDM8/SynVest-1 have not been reported in frontotemporal dementia yet. Several studies have used [^18^F]FDG PET to assess brain metabolism and interpreted changes as a marker of synaptic loss. Such metabolic change is disproportionate to atrophy (Bejanin *et al*., 2020), but is not a direct measure of synaptic density. The direct comparison between [^11^C]UCB-J and [^18^F]FDG PET in Alzheimer’s disease indicated a high correlation in medial temporal regions, but not elsewhere (Chen *et al*., 2021): measures of synaptic loss and hypometabolism may therefore provide complementary information about the underlying pathophysiology. In addition to [^18^F]FDG and [^11^C]UCB-J PET, synaptic function may be assessed by electro-/magneto-encephalography. For example, in Alzheimer’s disease, tau burden ([^18^F]flortaucipir) and synaptic loss ([^11^C]UCB-J) are both associated with changes in magnetoencephalography (Coomans *et al*., 2021). Fluidic markers of synaptic health are also emerging, including pentraxins (Van Der Ende *et al*., 2020), although these are currently largely confined to CSF-based rather than blood-based assays. In comparison to other techniques, [^11^C]UCB-J PET has excellent reproducibility which is advantageous for longitudinal and interventional studies.

Our study has several limitations. We acknowledge the relatively small sample size. However, the expected effect size in frontotemporal dementia was large (Cohen’s d>1), and power was estimated to be sufficient (β>0.8 for α<0.05). Moreover, we found consensus across different analytic methods: voxel-wise and ROI-based analyses, BP_ND_ values determined from data with and without partial-volume correction. The Bayesian tests confirmed that we had sufficient precision (analogous to power in frequentist tests) to support the alternate hypotheses (BFs > 3) with strong evidence from the eleven patients. The convergence over these statistical approaches mitigates against inadequate power and sample-dependant biases on the estimation of group differences and imaging-cognition associations. The replication of these findings with larger and multicentre clinical cohorts will nonetheless represent an important step to establish the generalizability of our results, and utility for clinical trials. Recruitment was based on clinical diagnosis, rather than neuropathology or genetics, but the clinical diagnosis was re-confirmed at serial clinical visits and has high clinico-pathological correlation with either FTLD-Tau or FTLD-TDP43. Given the novelty of the radiotracer, the cohort has been scanned recently (within 24 months of submission) meaning survival analyses and neuropathological confirmation are not yet possible. Future PET-to-autopsy studies will be needed to investigate the association between *in vivo* measures of synaptic density and neuropathology. Nonetheless, the concordance of partial-volume-corrected and non-corrected analyses already indicate that the synaptic loss we observe is not simply attributable to atrophy. To conclude, our study confirms that bvFTD is associated with significant and widespread frontotemporal loss of synapses, in proportion to disease severity. We suggest that [^11^C]UCB-J PET can facilitate the validation of preclinical models, inform models of human pathogenesis, and inform the design of new disease-modifying treatment strategies.

## Acknowledgements

We thank our participant volunteers and their caregivers for their participation in this study, the radiographers and radiochemists at the Wolfson Brain Imaging Centre, the FTD research nurses and clinical fellows at the Cambridge Centre for Frontotemporal Dementia and Related Disorders for their invaluable support. We also thank UCB Pharma for providing the precursor for [^11^C]UCB-J.

