## Supplementary Material for "Synaptic loss in behavioural variant frontotemporal dementia revealed by [^11^C]UCB-J PET"

### Supplemental Material

**Supplementary Table 1. Group comparisons of regional [$^{11}$C]UCB-J binding potential ($BP_{ND}$) with partial volume correction including aggregated regions.** P < 0.05 are highlighted in yellow, while p < 0.001 are highlighted in red.

| | $BP_{ND}$ | | | | Statistical parameters | | | |
|---|---|---|---|---|---|---|---|---|
| | **Mean CT** | **SD CT** | **Mean PT** | **SD PT** | **T value** | **Cohen's d** | **p** | **p FDR** |
| *Frontal L* | 2.87 | 0.22 | 2.22 | 0.33 | -5.99 | -2.33 | 0.0000 | 0.0001 |
| *Frontal R* | 2.90 | 0.21 | 2.28 | 0.31 | -6.03 | -2.33 | 0.0000 | 0.0001 |
| *Temporal L* | 2.57 | 0.24 | 1.96 | 0.30 | -6.08 | -2.28 | 0.0000 | 0.0001 |
| *Temporal R* | 2.62 | 0.22 | 2.03 | 0.32 | -5.63 | -2.17 | 0.0001 | 0.0002 |
| *Parietal L* | 3.04 | 0.24 | 2.54 | 0.29 | -5.03 | -1.89 | 0.0001 | 0.0003 |
| *Parietal R* | 3.01 | 0.22 | 2.53 | 0.31 | -4.71 | -1.81 | 0.0003 | 0.0004 |
| *Occipital L* | 2.90 | 0.28 | 2.56 | 0.33 | -3.06 | -1.14 | 0.0071 | 0.0075 |
| *Occipital R* | 2.89 | 0.25 | 2.51 | 0.35 | -3.19 | -1.23 | 0.0063 | 0.0071 |
| *Cingulate L* | 2.99 | 0.22 | 2.31 | 0.43 | -4.98 | -2.00 | 0.0003 | 0.0004 |
| *Cingulate R* | 2.94 | 0.24 | 2.24 | 0.37 | -5.80 | -2.26 | 0.0000 | 0.0002 |
| *Insula L* | 2.58 | 0.27 | 1.87 | 0.43 | -5.13 | -2.00 | 0.0002 | 0.0003 |
| *Insula R* | 2.69 | 0.26 | 1.79 | 0.39 | -6.91 | -2.68 | 0.0000 | 0.0001 |
| *Hippocampus L* | 1.84 | 0.27 | 1.36 | 0.53 | -2.84 | -1.14 | 0.0147 | 0.0147 |
| *Hippocampus R* | 1.84 | 0.28 | 1.17 | 0.39 | -5.12 | -1.96 | 0.0001 | 0.0003 |
| *Amygdala L* | 2.46 | 0.32 | 1.86 | 0.49 | -3.80 | -1.48 | 0.0020 | 0.0024 |
| *Amygdala R* | 2.50 | 0.30 | 1.57 | 0.55 | -5.26 | -2.09 | 0.0002 | 0.0003 |
| *Thalamus L* | 2.74 | 0.28 | 2.03 | 0.48 | -4.57 | -1.80 | 0.0005 | 0.0007 |
| *Thalamus R* | 2.86 | 0.31 | 1.86 | 0.79 | -4.07 | -1.67 | 0.0017 | 0.0022 |

Abbreviations: CT = controls, SD = standard deviation, PT = bvFTD patients, p = p value, FDR = false discovery rate correction, L = left, R = right.

**Supplementary Table 2. Group comparisons of regional [[11]C]UCB-J binding potential ($BP_{ND}$) with partial volume correction in Hammers atlas regions.** P < 0.05 are highlighted in yellow, while p < 0.001 are highlighted in red.

| | $BP_{ND}$ | | | | Statistical parameters | | | |
|---|---|---|---|---|---|---|---|---|
| | Mean CT | SD CT | Mean PT | SD PT | T value | Cohen's d | p | p FDR |
| *Hippocampus R* | 1.84 | 0.28 | 1.17 | 0.39 | -5.12 | -1.96 | 0.0001 | 0.0005 |
| *Hippocampus L* | 1.84 | 0.27 | 1.36 | 0.53 | -2.84 | -1.14 | 0.0147 | 0.0172 |
| *Amygdala R* | 2.50 | 0.30 | 1.57 | 0.55 | -5.26 | -2.09 | 0.0002 | 0.0005 |
| *Amygdala L* | 2.46 | 0.32 | 1.86 | 0.49 | -3.80 | -1.48 | 0.0020 | 0.0030 |
| *Ant_TL_med R* | 2.22 | 0.20 | 1.27 | 0.61 | -5.04 | -2.09 | 0.0004 | 0.0009 |
| *Ant_TL_med L* | 2.19 | 0.21 | 1.35 | 0.59 | -4.59 | -1.89 | 0.0008 | 0.0013 |
| *Ant_TL_inf Lat R* | 2.50 | 0.23 | 1.79 | 0.62 | -3.68 | -1.52 | 0.0035 | 0.0050 |
| *Ant_TL_inf Lat L* | 2.51 | 0.29 | 1.65 | 0.62 | -4.40 | -1.78 | 0.0009 | 0.0015 |
| *G_paraH_amb R* | 1.72 | 0.25 | 0.98 | 0.40 | -5.64 | -2.20 | 0.0001 | 0.0004 |
| *G_paraH_amb L* | 1.52 | 0.24 | 0.97 | 0.59 | -2.94 | -1.20 | 0.0128 | 0.0154 |
| *G_sup_temp_cent R* | 2.84 | 0.20 | 2.21 | 0.43 | -4.62 | -1.86 | 0.0006 | 0.0011 |
| *G_sup_temp_cent L* | 2.76 | 0.29 | 2.19 | 0.30 | -5.23 | -1.90 | 0.0001 | 0.0004 |
| *G_tem_midin R* | 2.66 | 0.24 | 1.89 | 0.44 | -5.50 | -2.19 | 0.0001 | 0.0005 |
| *G_tem_midin L* | 2.66 | 0.29 | 1.74 | 0.58 | -5.02 | -2.02 | 0.0003 | 0.0007 |
| *G_occtem La R* | 2.43 | 0.28 | 1.55 | 0.53 | -5.21 | -2.09 | 0.0002 | 0.0006 |
| *G_occtem La L* | 2.38 | 0.25 | 1.56 | 0.55 | -4.76 | -1.93 | 0.0005 | 0.0010 |
| *Insula L* | 2.58 | 0.27 | 1.87 | 0.43 | -5.13 | -2.00 | 0.0002 | 0.0005 |
| *Insula R* | 2.69 | 0.26 | 1.79 | 0.39 | -6.91 | -2.68 | 0.0000 | 0.0002 |
| *OL Rest Lat L* | 2.74 | 0.27 | 2.16 | 0.41 | -4.25 | -1.66 | 0.0008 | 0.0014 |
| *OL Rest Lat R* | 2.71 | 0.24 | 2.22 | 0.40 | -3.71 | -1.46 | 0.0025 | 0.0037 |
| *G_cing_ant_sup L* | 2.87 | 0.24 | 1.82 | 0.72 | -4.72 | -1.95 | 0.0006 | 0.0012 |
| *G_cing_ant_sup R* | 2.82 | 0.27 | 1.85 | 0.55 | -5.52 | -2.23 | 0.0001 | 0.0005 |
| *G_cing_post L* | 3.03 | 0.23 | 2.36 | 0.44 | -4.81 | -1.92 | 0.0004 | 0.0009 |
| *G_cing_post R* | 3.01 | 0.24 | 2.27 | 0.44 | -5.21 | -2.07 | 0.0002 | 0.0006 |
| *FL_mid_fr_G L* | 2.89 | 0.24 | 1.86 | 0.63 | -5.26 | -2.16 | 0.0002 | 0.0007 |
| *FL_mid_fr_G R* | 2.93 | 0.24 | 2.06 | 0.46 | -5.93 | -2.38 | 0.0001 | 0.0004 |
| *PosteriorTL L* | 2.69 | 0.26 | 1.94 | 0.34 | -6.49 | -2.47 | 0.0000 | 0.0002 |
| *PosteriorTL R* | 2.70 | 0.23 | 2.08 | 0.34 | -5.47 | -2.12 | 0.0001 | 0.0004 |
| *PL Rest L* | 2.84 | 0.25 | 2.09 | 0.40 | -5.77 | -2.25 | 0.0001 | 0.0004 |
| *PL Rest R* | 2.80 | 0.23 | 2.17 | 0.37 | -5.27 | -2.07 | 0.0001 | 0.0005 |
| *CaudateNucl L* | 2.99 | 0.34 | 2.55 | 0.44 | -2.94 | -1.12 | 0.0099 | 0.0124 |
| *CaudateNucl R* | 2.97 | 0.34 | 3.44 | 3.09 | 0.50 | 0.21 | 0.6284 | 0.6284 |
| *NuclAccumb L* | 3.84 | 0.33 | 3.03 | 0.77 | -3.31 | -1.35 | 0.0065 | 0.0086 |
| *NuclAccumb R* | 3.97 | 0.38 | 3.18 | 1.17 | -2.18 | -0.91 | 0.0520 | 0.0585 |
| *Putamen L* | 3.93 | 0.38 | 3.34 | 0.41 | -4.12 | -1.51 | 0.0007 | 0.0012 |
| *Putamen R* | 3.86 | 0.35 | 3.20 | 0.44 | -4.38 | -1.66 | 0.0005 | 0.0010 |
| *Thalamus L* | 2.74 | 0.28 | 2.03 | 0.48 | -4.57 | -1.80 | 0.0005 | 0.0010 |
| *Thalamus R* | 2.86 | 0.31 | 1.86 | 0.79 | -4.07 | -1.67 | 0.0017 | 0.0027 |
| *Pallidum L* | 1.71 | 0.27 | 1.27 | 0.28 | -4.34 | -1.59 | 0.0004 | 0.0009 |

| | | | | | | | | |
|---|---|---|---|---|---|---|---|---|
| *Pallidum R* | 1.96 | 0.25 | 1.57 | 0.29 | -3.93 | -1.46 | 0.0011 | 0.0018 |
| *FL_precen_G L* | 2.67 | 0.19 | 2.14 | 0.35 | -4.72 | -1.88 | 0.0004 | 0.0009 |
| *FL_precen_G R* | 2.67 | 0.19 | 2.20 | 0.29 | -4.97 | -1.92 | 0.0002 | 0.0006 |
| *FL_strai_G L* | 2.76 | 0.28 | 1.66 | 0.54 | -6.38 | -2.55 | 0.0000 | 0.0003 |
| *FL_strai_G R* | 2.71 | 0.25 | 1.61 | 0.62 | -5.75 | -2.35 | 0.0001 | 0.0005 |
| *FL_OFC_AOG L* | 2.73 | 0.23 | 1.98 | 0.54 | -4.22 | -1.80 | 0.0017 | 0.0027 |
| *FL_OFC_AOG R* | 2.78 | 0.23 | 2.21 | 0.89 | -2.07 | -0.87 | 0.0641 | 0.0692 |
| *FL_inf_fr_G L* | 2.82 | 0.24 | 1.81 | 0.59 | -5.51 | -2.25 | 0.0002 | 0.0005 |
| *FL_inf_fr_G R* | 2.83 | 0.21 | 1.95 | 0.58 | -4.92 | -2.03 | 0.0004 | 0.0009 |
| *FL_sup_fr_G L* | 2.84 | 0.25 | 1.93 | 0.49 | -5.85 | -2.35 | 0.0001 | 0.0004 |
| *FL_sup_fr_G R* | 2.87 | 0.26 | 2.04 | 0.42 | -6.00 | -2.36 | 0.0000 | 0.0004 |
| *PL_postce_G L* | 2.70 | 0.23 | 2.21 | 0.26 | -5.38 | -2.00 | 0.0001 | 0.0004 |
| *PL_postce_G R* | 2.68 | 0.22 | 2.21 | 0.25 | -5.26 | -1.96 | 0.0001 | 0.0004 |
| *PL_sup_pa_G L* | 3.12 | 0.25 | 2.40 | 0.33 | -6.49 | -2.46 | 0.0000 | 0.0002 |
| *PL_sup_pa_G R* | 3.08 | 0.23 | 2.45 | 0.29 | -6.24 | -2.36 | 0.0000 | 0.0002 |
| *OL Ling_G L* | 2.84 | 0.35 | 2.49 | 0.38 | -2.55 | -0.94 | 0.0204 | 0.0233 |
| *OL Ling_G R* | 2.96 | 0.30 | 2.55 | 0.38 | -3.15 | -1.19 | 0.0063 | 0.0085 |
| *OL_cuneus L* | 3.12 | 0.35 | 2.67 | 0.48 | -2.77 | -1.06 | 0.0144 | 0.0171 |
| *OL_cuneus R* | 3.14 | 0.33 | 2.69 | 0.37 | -3.47 | -1.29 | 0.0029 | 0.0042 |
| *FL_OFC_MOG L* | 2.63 | 0.28 | 1.62 | 0.62 | -5.16 | -2.09 | 0.0002 | 0.0007 |
| *FL_OFC_MOG R* | 2.66 | 0.30 | 1.70 | 0.67 | -4.61 | -1.87 | 0.0006 | 0.0012 |
| *FL_OFC LOG L* | 2.32 | 0.25 | 1.72 | 0.59 | -3.24 | -1.32 | 0.0073 | 0.0094 |
| *FL_OFC LOG R* | 2.43 | 0.25 | 1.72 | 0.70 | -3.26 | -1.34 | 0.0075 | 0.0095 |
| *FL_OFC_POG L* | 2.51 | 0.24 | 1.64 | 0.45 | -6.10 | -2.44 | 0.0000 | 0.0004 |
| *FL_OFC_POG R* | 2.53 | 0.23 | 1.55 | 0.61 | -5.18 | -2.13 | 0.0003 | 0.0007 |
| *Subgen_antCing L* | 2.73 | 0.31 | 1.57 | 1.13 | -3.35 | -1.40 | 0.0067 | 0.0088 |
| *Subgen_antCing R* | 2.57 | 0.38 | 1.52 | 1.01 | -3.36 | -1.38 | 0.0062 | 0.0085 |
| *Subcall_area L* | 3.11 | 0.57 | 1.89 | 3.17 | -1.27 | -0.54 | 0.2331 | 0.2452 |
| *Subcall_area R* | 2.92 | 0.59 | 2.20 | 3.68 | -0.65 | -0.28 | 0.5295 | 0.5429 |
| *Presubgen_antCing L* | 3.41 | 0.38 | 1.91 | 1.72 | -2.87 | -1.21 | 0.0159 | 0.0184 |
| *Presubgen_antCing R* | 3.26 | 0.42 | 2.02 | 1.35 | -2.98 | -1.24 | 0.0126 | 0.0154 |
| *G_sup_temp_ant L* | 2.25 | 0.26 | 2.03 | 1.31 | -0.56 | -0.24 | 0.5858 | 0.5931 |
| *G_sup_temp_ant R* | 2.34 | 0.26 | 1.84 | 0.39 | -3.86 | -1.50 | 0.0017 | 0.0027 |
| *Brainstem_mid* | 2.26 | 0.32 | 1.43 | 0.58 | -4.52 | -1.80 | 0.0006 | 0.0011 |
| *Brainstem_pon* | 0.96 | 0.15 | 0.75 | 0.17 | -3.56 | -1.32 | 0.0024 | 0.0035 |
| *Brainstem_med* | 0.67 | 0.15 | 0.45 | 0.20 | -3.20 | -1.22 | 0.0059 | 0.0082 |
| *Cerebellum_gm R* | 2.13 | 0.21 | 1.72 | 0.15 | -6.63 | -2.24 | 0.0000 | 0.0000 |
| *Cerebellum_gm L* | 1.44 | 0.73 | 1.71 | 0.19 | 1.73 | 0.51 | 0.0934 | 0.0996 |
| *Cerebellum_wm R* | 1.92 | 1.18 | 1.22 | 0.87 | -2.00 | -0.68 | 0.0563 | 0.0624 |
| *Cerebellum_wm L* | 1.67 | 0.74 | 1.86 | 0.82 | 0.68 | 0.25 | 0.5029 | 0.5222 |
| *Cerebellum_dentate L* | 1.05 | 0.16 | 0.87 | 0.28 | -2.08 | -0.82 | 0.0578 | 0.0633 |
| *Cerebellum_dentate R* | 1.77 | 0.66 | 0.82 | 0.24 | -6.29 | -1.91 | 0.0000 | 0.0000 |

Abbreviations: CT = controls, SD = standard deviation, PT = bvFTD patients, p = p value, FDR = false discovery rate correction, L = left, R = right.

**Supplementary Table 3. Group comparisons of regional [$^{11}$C]UCB-J binding potential ($BP_{ND}$) without partial volume correction in Hammers atlas regions.** P < 0.05 are highlighted in yellow, while p < 0.001 are highlighted in red.

| | $BP_{ND}$ | | | | Statistical parameters | | | |
|---|---|---|---|---|---|---|---|---|
| | Mean CT | SD CT | Mean PT | SD PT | T value | Cohen's d | p | p FDR |
| *Hippocampus R* | 1.90 | 0.22 | 1.15 | 0.29 | -7.74 | -2.94 | 0.0000 | 0.0000 |
| *Hippocampus L* | 1.89 | 0.21 | 1.27 | 0.36 | -5.39 | -2.13 | 0.0001 | 0.0002 |
| *Amygdala R* | 2.34 | 0.24 | 1.48 | 0.45 | -6.02 | -2.40 | 0.0000 | 0.0001 |
| *Amygdala L* | 2.32 | 0.25 | 1.67 | 0.42 | -4.74 | -1.87 | 0.0004 | 0.0005 |
| *Ant_TL_med R* | 1.92 | 0.19 | 1.18 | 0.40 | -5.88 | -2.38 | 0.0001 | 0.0002 |
| *Ant_TL_med L* | 1.90 | 0.20 | 1.20 | 0.43 | -5.09 | -2.06 | 0.0003 | 0.0004 |
| *Ant_TL_inf Lat R* | 2.15 | 0.21 | 1.47 | 0.42 | -5.05 | -2.03 | 0.0003 | 0.0004 |
| *Ant_TL_inf Lat L* | 2.20 | 0.27 | 1.43 | 0.46 | -5.15 | -2.03 | 0.0002 | 0.0003 |
| *G_paraH_amb R* | 1.73 | 0.21 | 1.08 | 0.28 | -6.87 | -2.63 | 0.0000 | 0.0000 |
| *G_paraH_amb L* | 1.59 | 0.20 | 1.07 | 0.33 | -4.78 | -1.88 | 0.0003 | 0.0004 |
| *G_sup_temp_cent R* | 2.03 | 0.17 | 1.48 | 0.24 | -6.89 | -2.66 | 0.0000 | 0.0000 |
| *G_sup_temp_cent L* | 1.97 | 0.22 | 1.45 | 0.24 | -5.96 | -2.20 | 0.0000 | 0.0001 |
| *G_tem_midin R* | 2.08 | 0.21 | 1.46 | 0.32 | -5.99 | -2.33 | 0.0000 | 0.0001 |
| *G_tem_midin L* | 2.06 | 0.24 | 1.36 | 0.40 | -5.32 | -2.09 | 0.0001 | 0.0002 |
| *G_occtem La R* | 2.14 | 0.24 | 1.40 | 0.43 | -5.37 | -2.13 | 0.0001 | 0.0002 |
| *G_occtem La L* | 2.11 | 0.21 | 1.41 | 0.42 | -5.26 | -2.12 | 0.0002 | 0.0003 |
| *Insula L* | 1.88 | 0.18 | 1.36 | 0.32 | -5.05 | -2.00 | 0.0002 | 0.0003 |
| *Insula R* | 2.01 | 0.19 | 1.35 | 0.29 | -7.05 | -2.75 | 0.0000 | 0.0000 |
| *OL Rest Lat L* | 1.95 | 0.20 | 1.52 | 0.27 | -4.84 | -1.85 | 0.0002 | 0.0003 |
| *OL Rest Lat R* | 1.94 | 0.18 | 1.57 | 0.26 | -4.22 | -1.63 | 0.0008 | 0.0009 |
| *G_cing_ant_sup L* | 2.16 | 0.21 | 1.33 | 0.47 | -5.68 | -2.30 | 0.0001 | 0.0002 |
| *G_cing_ant_sup R* | 2.05 | 0.23 | 1.33 | 0.38 | -5.90 | -2.32 | 0.0000 | 0.0001 |
| *G_cing_post L* | 2.32 | 0.21 | 1.67 | 0.36 | -5.54 | -2.18 | 0.0001 | 0.0002 |
| *G_cing_post R* | 2.26 | 0.21 | 1.61 | 0.30 | -6.56 | -2.53 | 0.0000 | 0.0001 |
| *FL_mid_fr_G L* | 1.48 | 0.17 | 0.88 | 0.31 | -5.94 | -2.36 | 0.0001 | 0.0001 |
| *FL_mid_fr_G R* | 1.58 | 0.18 | 1.05 | 0.28 | -5.78 | -2.26 | 0.0001 | 0.0001 |
| *PosteriorTL L* | 1.89 | 0.21 | 1.34 | 0.26 | -6.20 | -2.33 | 0.0000 | 0.0001 |
| *PosteriorTL R* | 1.92 | 0.19 | 1.46 | 0.24 | -5.63 | -2.14 | 0.0000 | 0.0001 |
| *PL Rest L* | 1.86 | 0.20 | 1.29 | 0.29 | -5.96 | -2.29 | 0.0000 | 0.0001 |
| *PL Rest R* | 1.89 | 0.18 | 1.42 | 0.25 | -5.64 | -2.17 | 0.0001 | 0.0001 |
| *CaudateNucl L* | 2.35 | 0.25 | 1.51 | 0.55 | -4.80 | -1.95 | 0.0005 | 0.0006 |
| *CaudateNucl R* | 2.30 | 0.26 | 1.44 | 0.49 | -5.46 | -2.18 | 0.0001 | 0.0002 |
| *NuclAccumb L* | 3.01 | 0.25 | 2.16 | 0.51 | -5.27 | -2.13 | 0.0002 | 0.0003 |
| *NuclAccumb R* | 3.03 | 0.25 | 2.12 | 0.52 | -5.51 | -2.22 | 0.0001 | 0.0002 |
| *Putamen L* | 3.12 | 0.30 | 2.49 | 0.39 | -4.77 | -1.81 | 0.0002 | 0.0003 |
| *Putamen R* | 3.19 | 0.29 | 2.55 | 0.40 | -4.83 | -1.85 | 0.0002 | 0.0003 |
| *Thalamus L* | 2.12 | 0.19 | 1.69 | 0.28 | -4.70 | -1.81 | 0.0003 | 0.0004 |
| *Thalamus R* | 2.11 | 0.19 | 1.67 | 0.24 | -5.34 | -2.01 | 0.0001 | 0.0001 |
| *Pallidum L* | 1.33 | 0.20 | 1.04 | 0.19 | -4.13 | -1.48 | 0.0005 | 0.0006 |
| *Pallidum R* | 1.54 | 0.19 | 1.27 | 0.19 | -3.98 | -1.43 | 0.0007 | 0.0009 |

| | | | | | | | | |
|---|---|---|---|---|---|---|---|---|
| *FL_precen_G L* | 1.39 | 0.11 | 1.08 | 0.18 | -5.13 | -2.01 | 0.0002 | 0.0003 |
| *FL_precen_G R* | 1.43 | 0.12 | 1.15 | 0.16 | -5.21 | -2.00 | 0.0001 | 0.0002 |
| *FL_strai_G L* | 2.44 | 0.26 | 1.45 | 0.39 | -7.57 | -2.94 | 0.0000 | 0.0000 |
| *FL_strai_G R* | 2.38 | 0.23 | 1.44 | 0.39 | -7.53 | -2.97 | 0.0000 | 0.0000 |
| *FL_OFC_AOG L* | 2.00 | 0.18 | 1.40 | 0.32 | -5.91 | -2.34 | 0.0001 | 0.0001 |
| *FL_OFC_AOG R* | 2.07 | 0.16 | 1.46 | 0.26 | -7.13 | -2.79 | 0.0000 | 0.0000 |
| *FL_inf_fr_G L* | 2.09 | 0.20 | 1.32 | 0.34 | -7.09 | -2.80 | 0.0000 | 0.0000 |
| *FL_inf_fr_G R* | 2.15 | 0.17 | 1.45 | 0.30 | -7.14 | -2.82 | 0.0000 | 0.0000 |
| *FL_sup_fr_G L* | 1.76 | 0.19 | 1.14 | 0.31 | -6.17 | -2.42 | 0.0000 | 0.0001 |
| *FL_sup_fr_G R* | 1.76 | 0.19 | 1.19 | 0.26 | -6.65 | -2.54 | 0.0000 | 0.0000 |
| *PL_postce_G L* | 1.54 | 0.14 | 1.18 | 0.21 | -5.20 | -2.01 | 0.0001 | 0.0002 |
| *PL_postce_G R* | 1.54 | 0.14 | 1.22 | 0.17 | -5.47 | -2.05 | 0.0000 | 0.0001 |
| *PL_sup_pa_G L* | 1.80 | 0.17 | 1.30 | 0.20 | -7.21 | -2.70 | 0.0000 | 0.0000 |
| *PL_sup_pa_G R* | 1.81 | 0.16 | 1.34 | 0.19 | -7.27 | -2.69 | 0.0000 | 0.0000 |
| *OL Ling_G L* | 2.42 | 0.25 | 2.01 | 0.34 | -3.58 | -1.37 | 0.0027 | 0.0030 |
| *OL Ling_G R* | 2.43 | 0.23 | 2.03 | 0.32 | -3.73 | -1.43 | 0.0021 | 0.0023 |
| *OL_cuneus L* | 2.51 | 0.25 | 2.00 | 0.38 | -4.04 | -1.57 | 0.0012 | 0.0014 |
| *OL_cuneus R* | 2.50 | 0.27 | 2.04 | 0.38 | -3.63 | -1.40 | 0.0026 | 0.0028 |
| *FL_OFC_MOG L* | 2.05 | 0.20 | 1.38 | 0.32 | -6.46 | -2.52 | 0.0000 | 0.0001 |
| *FL_OFC_MOG R* | 2.05 | 0.21 | 1.40 | 0.31 | -6.27 | -2.42 | 0.0000 | 0.0001 |
| *FL_OFC LOG L* | 1.91 | 0.20 | 1.39 | 0.28 | -5.57 | -2.14 | 0.0001 | 0.0001 |
| *FL_OFC LOG R* | 1.98 | 0.19 | 1.45 | 0.34 | -4.90 | -1.94 | 0.0003 | 0.0004 |
| *FL_OFC_POG L* | 2.02 | 0.20 | 1.40 | 0.26 | -6.99 | -2.66 | 0.0000 | 0.0000 |
| *FL_OFC_POG R* | 2.07 | 0.18 | 1.36 | 0.33 | -6.73 | -2.68 | 0.0000 | 0.0001 |
| *Subgen_antCing L* | 1.90 | 0.26 | 1.19 | 0.39 | -5.57 | -2.16 | 0.0001 | 0.0001 |
| *Subgen_antCing R* | 1.65 | 0.24 | 1.03 | 0.37 | -5.12 | -1.99 | 0.0002 | 0.0002 |
| *Subcall_area L* | 2.22 | 0.27 | 1.31 | 0.44 | -6.30 | -2.47 | 0.0000 | 0.0001 |
| *Subcall_area R* | 2.24 | 0.28 | 1.29 | 0.48 | -6.11 | -2.41 | 0.0000 | 0.0001 |
| *Presubgen_antCing L* | 2.65 | 0.30 | 1.63 | 0.50 | -6.27 | -2.47 | 0.0000 | 0.0001 |
| *Presubgen_antCing R* | 2.43 | 0.27 | 1.53 | 0.53 | -5.36 | -2.15 | 0.0002 | 0.0002 |
| *G_sup_temp_ant L* | 1.99 | 0.23 | 1.36 | 0.37 | -5.17 | -2.02 | 0.0002 | 0.0002 |
| *G_sup_temp_ant R* | 2.06 | 0.23 | 1.48 | 0.33 | -5.23 | -2.02 | 0.0001 | 0.0002 |
| *Brainstem_mid* | 1.07 | 0.12 | 0.84 | 0.22 | -3.27 | -1.31 | 0.0063 | 0.0068 |
| *Brainstem_pon* | 0.91 | 0.14 | 0.73 | 0.15 | -3.51 | -1.29 | 0.0025 | 0.0027 |
| *Brainstem_med* | 0.71 | 0.15 | 0.53 | 0.19 | -2.75 | -1.05 | 0.0146 | 0.0146 |
| *Cerebellum_gm R* | 2.02 | 0.20 | 1.59 | 0.17 | -6.55 | -2.30 | 0.0000 | 0.0000 |
| *Cerebellum_gm L* | 2.00 | 0.21 | 1.58 | 0.20 | -5.73 | -2.05 | 0.0000 | 0.0001 |
| *Cerebellum_wm R* | 0.81 | 0.12 | 0.70 | 0.10 | -2.75 | -0.95 | 0.0111 | 0.0116 |
| *Cerebellum_wm L* | 0.81 | 0.12 | 0.69 | 0.11 | -2.83 | -1.02 | 0.0103 | 0.0109 |
| *Cerebellum_dentate L* | 0.99 | 0.13 | 0.81 | 0.20 | -2.86 | -1.10 | 0.0123 | 0.0126 |
| *Cerebellum_dentate R* | 0.88 | 0.12 | 0.74 | 0.16 | -2.77 | -1.06 | 0.0144 | 0.0146 |

Abbreviations: CT = controls, SD = standard deviation, PT = bvFTD patients, p = p value, FDR = false discovery rate correction, L = left, R = right.

**Supplementary Table 4. Spearman correlations in the bvFTD group between cognitive scores and $[^{11}C]$UCB-J binding potential ($BP_{ND}$) in aggregated cortical regions of interest.** Spearman rho (r) is reported alongside p value and Bayesian factor (BF), with * denoting a p value < 0.05.

| REGION | ACE-R | ATT/OR | MEM | FLUE | LANG | VISUO | IFS | IFS WM | MMSE | FTD-RS % |
|---|---|---|---|---|---|---|---|---|---|---|
| **Frontal L** | r=0.791 (0.002)* BF=56.2 | r=0.717 (0.007)* BF=21.3 | r=0.653 (0.015)* BF=7.9 | r=0.954 (< .001)* BF=4720.6 | r=0.795 (0.002)* BF=52.5 | r=0.816 (0.001)* BF=71.4 | r=0.754 (0.006)* BF=29.5 | r=0.74 (0.007)* BF=23.4 | r=0.736 (0.005)* BF=20.6 | r=-0.055 (0.446) BF=0.4 |
| **Frontal R** | r=0.118 (0.367) BF=0.4 | r=0.028 (0.467) BF=0.3 | r=0.027 (0.468) BF=0.4 | r=0.396 (0.114) BF=1.2 | r=0.354 (0.143) BF=1 | r=0.06 (0.431) BF=0.3 | r=0.17 (0.319) BF=0.7 | r=0.177 (0.312) BF=0.7 | r=0.133 (0.348) BF=0.4 | r=-0.382 (0.139) BF=0.2 |
| **Temporal L** | r=0.445 (0.086) BF=1.7 | r=0.422 (0.098) BF=1.5 | r=0.443 (0.086) BF=1.8 | r=0.691 (0.009)* BF=12.8 | r=0.612 (0.023)* BF=8.2 | r=0.581 (0.031)* BF=4.3 | r=0.547 (0.051) BF=3.4 | r=0.495 (0.073) BF=2.5 | r=0.382 (0.123) BF=1 | r=-0.042 (0.459) BF=0.4 |
| **Temporal R** | r=-0.245 (0.774) BF=0.2 | r=-0.244 (0.765) BF=0.2 | r=-0.174 (0.695) BF=0.2 | r=0.06 (0.431) BF=0.4 | r=0.032 (0.463) BF=0.4 | r=-0.088 (0.601) BF=0.2 | r=0.018 (0.48) BF=0.4 | r=-0.012 (0.513) BF=0.4 | r=-0.271 (0.79) BF=0.2 | r=-0.139 (0.354) BF=0.3 |
| **Cingulate L** | r=0.700 (0.008)* BF=10.7 | r=0.619 (0.021)* BF=4.9 | r=0.589 (0.028)* BF=3.6 | r=0.82 (< .001)* BF=46.5 | r=0.814 (0.001)* BF=88.9 | r=0.604 (0.025)* BF=6.1 | r=0.62 (0.028)* BF=2.3 | r=0.581 (0.039)* BF=1.9 | r=0.713 (0.007)* BF=9.1 | r=-0.067 (0.432) BF=0.5 |
| **Cingulate R** | r=0.073 (0.419) BF=0.5 | r=0.141 (0.34) BF=0.5 | r=-0.032 (0.537) BF=0.3 | r=0.3 (0.185) BF=0.9 | r=0.221 (0.257) BF=0.6 | r=0.041 (0.452) BF=0.3 | r=0.134 (0.356) BF=0.7 | r=0.116 (0.375) BF=0.8 | r=0.175 (0.304) BF=0.6 | r=-0.115 (0.379) BF=0.4 |
| **Insula L** | r=0.264 (0.217) BF=0.9 | r=0.314 (0.174) BF=0.9 | r=0.237 (0.241) BF=0.7 | r=0.562 (0.036)* BF=4.2 | r=0.446 (0.085) BF=2.4 | r=0.452 (0.082) BF=1.7 | r=0.48 (0.08) BF=3.8 | r=0.428 (0.109) BF=2.8 | r=0.257 (0.222) BF=0.7 | r=-0.273 (0.224) BF=0.2 |
| **Insula R** | r=-0.136 (0.663) BF=0.3 | r=-0.028 (0.533) BF=0.3 | r=-0.137 (0.656) BF=0.3 | r=0.138 (0.343) BF=0.5 | r=0.078 (0.41) BF=0.4 | r=-0.065 (0.575) BF=0.3 | r=0.061 (0.434) BF=0.6 | r=0.037 (0.46) BF=0.6 | r=-0.041 (0.548) BF=0.3 | r=-0.285 (0.214) BF=0.2 |
| **Parietal L** | r=0.591 (0.028)* BF=3.9 | r=0.351 (0.145) BF=1.3 | r=0.616 (0.022)* BF=5.7 | r=0.627 (0.02)* BF=5.9 | r=0.809 (0.001)* BF=30.5 | r=0.461 (0.077) BF=1.9 | r=0.632 (0.025)* BF=0.8 | r=0.575 (0.041)* BF=0.7 | r=0.377 (0.127) BF=1.2 | r=-0.042 (0.459) BF=1.8 |
| **Parietal R** | r=-0.109 (0.633) BF=0.3 | r=-0.216 (0.738) BF=0.3 | r=-0.027 (0.532) BF=0.4 | r=-0.157 (0.677) BF=0.3 | r=0.143 (0.338) BF=0.5 | r=-0.23 (0.752) BF=0.2 | r=-0.14 (0.65) BF=0.2 | r=-0.22 (0.729) BF=0.2 | r=-0.216 (0.738) BF=0.3 | r=0.079 (0.594) BF=3 |
| **Occipital L** | r=-0.255 (0.783) BF=0.2 | r=-0.145 (0.665) BF=0.2 | r=-0.251 (0.772) BF=0.2 | r=-0.051 (0.559) BF=0.3 | r=-0.299 (0.814) BF=0.2 | r=0.000 (0.5) BF=0.3 | r=-0.201 (0.711) BF=0.3 | r=-0.196 (0.706) BF=0.3 | r=-0.28 (0.798) BF=0.2 | r=0.091 (0.607) BF=0.5 |
| **Occipital R** | r=-0.355 (0.863) BF=0.2 | r=-0.248 (0.769) BF=0.2 | r=-0.315 (0.827) BF=0.2 | r=-0.147 (0.667) BF=0.2 | r=-0.34 (0.847) BF=0.2 | r=-0.138 (0.657) BF=0.3 | r=-0.292 (0.793) BF=0.2 | r=-0.287 (0.79) BF=0.3 | r=-0.372 (0.87) BF=0.2 | r=0.236 (0.754) BF=0.6 |

Abbreviations: ACE-R = Addenbrooke's cognitive examination revised; ATT/OR= attention/orientation score; MEM = memory; FLUE = fluency; LANG = language; VISUO= visuospatial; IFS = INECO frontal screening; MMSE = mini-mental state examination; WM = working memory; FTD-RS = frontotemporal dementia rating scale; L = left; R = right.
